# An unsupervised machine learning approach to predict recovery from traumatic spinal cord injury

**DOI:** 10.1101/2023.09.26.23295361

**Authors:** Sarah C. Brüningk, Lucie Bourguignon, Louis P. Lukas, EMSCI study group, Doris Maier, Rainer Abel, Norbert Weidner, Rüdiger Rupp, Fred Geisler, John L.K. Kramer, James Guest, Armin Curt, Catherine R. Jutzeler

**Affiliations:** Department of Health Sciences and Technology (D-HEST), ETH Zurich, Universitätstrasse 2, 8092 Zürich, Switzerland; Spinal Cord Injury Center, University Hospital Balgrist, University of Zurich, Switzerland; Spinal Cord Injury Center, Trauma Center Murnau, Murnau, Germany; Spinal Cord Injury Center, Klinikum Bayreuth, Bayreuth, Germany; Spinal Cord Injury Center, Heidelberg University Hospital, Heidelberg, Germany; University of Saskatchewan, Saskatchewan, Canada; International Collaboration on Repair Discoveries (ICORD), University of British Columbia, Canada; Djavad Mowafaghian Centre for Brain Health, University of British Columbia, Canada; Department of Anesthesiology, Pharmacology, and Therapeutics, Faculty of Medicine, and University of British Columbia, Canada; Hugill Centre for Anesthesia, University of British Columbia, Canada; The Miami Project to Cure Paralysis, Miller School of Medicine, The University of Miami, Miami, United States; Department of Neurological Surgery, Miller School of Medicine, The University of Miami, Miami, United States; Schulthess Clinic, Zurich, Switzerland

**Keywords:** K-nearest neighbour, kNN, spinal cord injury, machine learning, recovery prediction

## Abstract

**Background:** Neurological and functional recovery after traumatic spinal cord injury (SCI) is highly heterogeneous, challenging outcome predictions in rehabilitation and clinical trials. We propose k-nearest neighbour (k-NN) matching as a data-driven, interpretable solution.

**Methods:** This study used acute-phase International Standards for Neurological Classification of SCI exams to forecast 6-month recovery motor function as primary evaluation endpoint. Secondary endpoints included severity grade improvement, independent walking, and self-care ability. Different similarity metrics were explored for NN matching within 1267 patients from the European Multicenter Study about Spinal Cord Injury before validation in 411 patients from the Sygen trial.

**Results:** We obtained a population-wide root-mean-squared error (RMSE) in motor score sequence of 0.76(0.14, 2.77) and competitive functional score predictions (AUC_walker_=0.92, AUC_self-carer_=0.83). The validation cohort showed comparable results (RMSE = 0.75(0.13, 2.57), AUC_walker_=0.92). Prediction performance in AIS grade B and C patients (∼30%) showed the largest deviations from true recovery scores, in line with large SCI heterogeneity.

**Conclusions:** Our approach provides detailed predictions of neurological and functional recovery based on a highly interpretable unsupervised machine learning concept. The k-NN matching strategy further enables the integration of historical control data into the evaluation of clinical trials and provides a data-driven digital twin for recovery trajectory exploration.

## 1. Background

Traumatic spinal cord injury (SCI) greatly impacts the quality of life of affected individuals due to an impairment of sensorimotor function. (1,2) Patients and their families have critical questions regarding their prognosis of recovery and probable long-term disability to plan for their health-related, financial, and social future. Personalised outcome predictions could provide a more specific perspective and help guide physicians in their rehabilitation planning. However, accurate prediction of sensorimotor recovery from traumatic SCI for an individual is challenging as injury and recovery patterns of traumatic SCI are greatly heterogeneous (3,4). The International Standards for Neurological Classification of SCI (ISNCSCI) exam assessing sensorimotor functions in a standardised way represents the global clinical gold standard to determine the neurological level and injury severity. This exam assesses light touch, pinprick and motor function as integer scores from 0 to 2 (sensation) or 5, respectively. The American Spinal Injury Association (ASIA) impairment scale (AIS) further gives a notion of the overall injury severity. Sensory and motor scores, the impairment scale grade, together with the relevant patient age, sex, and neurological level of injury (NLI) mainly provide the current basis for recovery prognosis.(5–7) Although some personalised biomarkers of recovery extracted from magnetic resonance imaging or injury-derived proteins have also been reported (8–12), recovery estimates are mainly determined on the basis of changes in values of ordinal scales such as the ISNCSCI on a group level. The current state-of-the-art in the field is a logistic regression model by van Middendrop et al. (13) that provided a prediction of an individual’s walking ability. Despite this success, current prognostic models are greatly limited in terms of flexibility to predict multiple functional abilities and neurological recovery which may be valuable to assess the full clinical picture of a patient. To the best of our knowledge, there currently exists no publicly available tool providing a more holistic recovery predictions covering several aspects of the recovery process. At the same time, understanding the prediction process and the confounding variables leading to a specific outcome prediction are key and often not easily accessible for the individual. Similarly, a notion of prediction uncertainty given input data variability should be provided in light of medical assessment data such as the ISNCSCI (14).

Despite numerous clinical trials over the last decades, the range of effective treatments to ameliorate traumatic damage to the spinal cord is limited. The heterogeneity of outcomes and the scarcity of reliable outcome predictors have likely contributed to the failure of SCI clinical trials, where, due to its low incidence (8.0 to 246.0 cases per million inhabitants per year), recruitment of participants is highly challenging (15). A potential mitigation strategy may be to use historic control cohorts as an extension to placebo-controlled trial designs by maximising the number of patients receiving the investigational therapy. This approach has been adopted by the Nogo Inhibition in Spinal Cord Injury (NISCI) trial (clinicaltrials.gov #NCT03935321), a current phase-II study that followed a phase-I evaluation by Kucher *et al*.(16). NISCI randomises patients at a ratio of 1:2 into placebo and treatment arms while accounting for the smaller randomised control group through historical controls. Integral to the success of such advanced trial designs is a reliable means of identifying suitable historic controls. Currently, a comprehensive comparison of different ways to match historical controls is also missing.

In this study, we define “historic neighbours” as the closest match(es) to a patient within a reference database in the acute phase of SCI (see **Figure 1A**) as a purely data-driven, unsupervised process. In contrast to other data-driven approaches such as supervised machine or deep learning (17–20), k-nearest neighbour matching (21) provides an inherently interpretable solution that is flexible to predict multiple endpoints simultaneously. The transparency of the prediction pipeline is essential for high-stakes clinical decision-making to optimise the resource investment in clinical trials. As a so-called lazy learner algorithm, k-NN, does not involve a specific training phase but uses the reference data upon inference to identify “similarity” between the sample of interest and the reference pool. The relevant similarity metric needs to be carefully chosen to reflect the clinical hallmarks of the task at hand.

**Figure 1:**
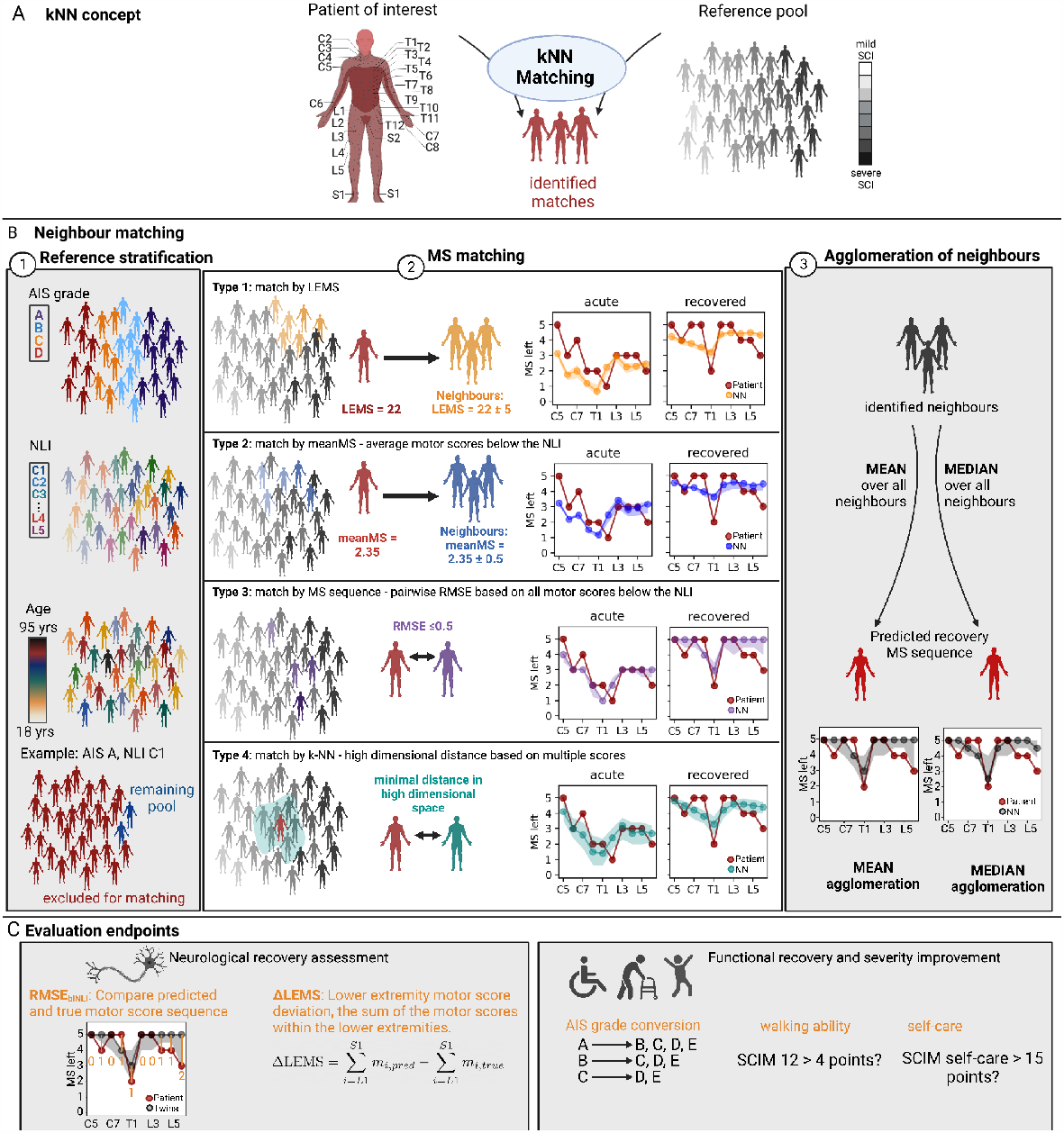
Study overview. A: Simplified schematic of the pipeline used in this study. Patients of interest (red) are characterized by the ISNCSCI exam and matched to neighbours according to a predefined matching algorithm from a large reference pool (here the EMSCI database, black). SCI severity is here shown as an abstract colour scale only. B: Overview of the investigated nearest neighbour matching methods. Matching starts with an optional stratification of the reference pool (1) to match AIS grade, NLI and/or patient age (within +/- 10 years for each patient, not by age categories), and sex to the patient of interest, followed by the identification of neighbours based on one of four types of motor function pattern quantifications (2). MS matching is based on one of four types outlined in **Table 1**. For each MS matching type results for a representative tetraplegic AIS C patient are shown. The true (red) and neighbour MS sequences are shown as graphs, whose y-scale is the MS quantity, and the successive myotome location is encoded in the x-direction. Lines emphasise the serial nature of these scores. Uncertainty bands stem from bootstrapping. The final recovery prediction constitutes an agglomeration of the recovery phase MSs of all identified neighbours by mean or median (3). C: Evaluation endpoints assessed including scores quantifying neurological and functional recovery. *Abbreviations*: SCI: spinal cord injury, AIS: American Spinal Injury Association impairment scale, NLI: neurological level of injury, LEMS: lower extremity motor score, RMSE_blNLI_: root-mean-squared error below the NLI, MS: motor score, meanMS: mean motor score, kNN: k-nearest neighbour, SCIM: spinal cord independence measure.

The objective of this study was to benchmark different ways to match patients with SCI to historic cohorts (i.e. the nearest neighbours) and quantify the degree to which these models provide detailed, personalised recovery predictions. The central hypothesis of this study is that kNN regression would be superior to patient stratification by clinical and demographic features alone. The mechanistic rationale for this is that several features of the retained motor and sensory function influence recovery implying that the full motor function pattern needs to be accounted for upon neighbour matching. Particularly, we score both neurological recovery (at the myotome level) and functional ability to provide a full picture of clinical SCI recovery. Where possible we benchmark our approach against the current state-of-the-art, i.e. a logistic regression for walking ability prediction and a regularised linear (Ridge) regression for the prediction of segmental motos scores. Importantly, by making the analysis code publicly available it is possible to freely use the new paradigms for clinical trial design and personalised predictions.

## 2. Methods

**Figure 1** summarizes the workflow of this study.

### 2.1. Included Data

We based the analysis on the European Multicenter Study about Spinal Cord Injury (clinicaltrials.gov #NCT01571531), comprising over 5000 patients as of April 2021. The EMSCI study is performed in accordance with the Declaration of Helsinki and approved by all contributing institutional review boards. We validate our findings against an independent cohort from the Sygen trial(22).

#### Patient inclusion criteria

Included traumatic SCI patients were required to have complete ISNCSCI (23,24) assessments and AIS grades in the very acute (within two weeks of injury) and the recovery phase (∼26 weeks, 150-186 days). We excluded patients with an NLI in the sacral segments and those displaying notable deterioration in motor function (> 2 points per score). Given no treatment effect was demonstrated by the Sygen trial we pooled control and intervention groups (22,25). Despite significant changes in patient management (26,27), we included all historic patient data given no proven improvement over time(28). This led to 1267 patients from the EMSCI database (**Table 1, Figure S1**) being identified as a reference pool and assessment of the kNN by leave-one-out cross-validation. Sygen patients were matched to the EMSCI reference pool.

**Table 1:**
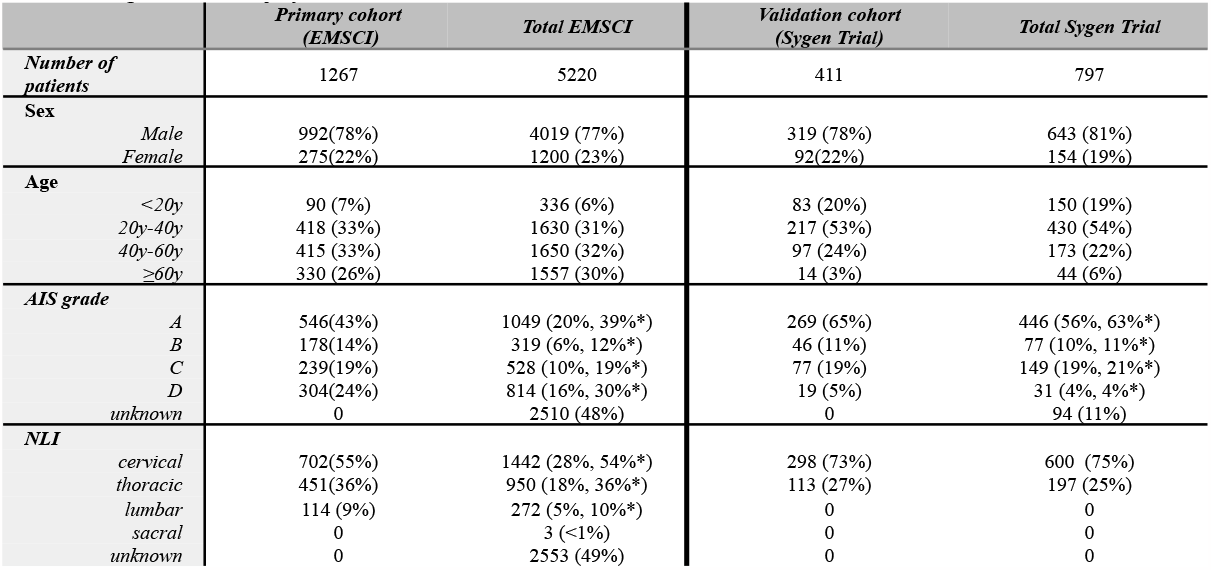
Patient characteristics for the cohorts included in this analysis. Patients were selected from the relevant databases based on the assessment of motor and sensory function at both the acute injury phase and 26 weeks after the injury occurred. Any patients that deteriorated by more than one MS point within any myotome were excluded. *percentages excluding unknowns. *Abbreviations:* EMSCI: European Multicenter Study about Spinal Cord Injury; AIS: American Spinal Injury Association Impairment Scale; NLI: Neurological level of injury.

#### *Included variables for neighbour matching* (2-week assessments)

All ISNCSCI scores, including motor (MS) and sensory scores (SS, i.e. light touch (LTS) and pinprick (PPS)) and the presence or absence of deep anal pressure (DAP) sensation and the ability for any voluntary anal contraction (VAC) were used. For optional patient matching was performed based on the NLI, patient demographics (age, sex), the AIS grade, and lower extremity motos score (LEMS).

#### *Evaluation endpoints* (26-week assessments)

**Figure 1C** gives an overview of the neurological and functional evaluation endpoints used. The primary evaluation endpoint was neurological recovery of motor function quantified as root mean squared errors below the NLI between the true and predicted MSs for an individual. Secondary endpoints included differences in LEMS (ΔLEMS), AIS grade conversion, and the ability to walk independently(13) or to care for oneself. For the latter, we binarize the Spinal Cord Independence Measure (SCIM) subscores (#1-4) assessing self-care into self-carers (SCIM_1-4_ ≥ 15, i.e reaching at least 75%) and dependent patients (SCIM_1-4_ <15). During the EMSCI trial, different versions of the SCIM (II(29) and III(30)) were used which were pooled and available for 1189 of the 1267 patients. For Sygen patients, SCIM was not assessed. Instead Benzel et. al.’s modified Japanese Orthopaedic Association Scale (31) with equivalent cutoffs to predict walking ability was used. Self-care potential was not assessed.

### 2.2. Similarity metrics and optimal hyperparameter selection

All of the described concepts were implemented in *python* (version 3.8). **Figure 1B** outlines the three-step procedure followed for neighbour matching and prediction calculation: Reference pool stratification (step 1), MS matching (step 2), neighbour agglomeration (step 3). Step 1 comprises an optional stratification by variables not representing the motor function pattern (NLI, AIS grade, sex, age +/- 10-year bracket) to account for biases. I.e. all retained reference patients matched the patient of interest for these variables. In step 2 the sensorimotor function of the patient of interest was matched based on one of four similarity metrics summarised in **Table 2** matching either LEMS (+/- 5 points, Type 1), or the mean MS below the NLI (+/- 0.5 points, Type 2), or achieved an RMSE between the acute-phase MS below the NLI with that of the patient of interest below 0.5 (Type 3), or were identified as kNN (with k in [1,3,10,20]) based on either all MS, or both MS and SS (LTS, PPS). The recovery-phase outcome variables of all identified neighbours are agglomerated by either mean or median in the final step to arrive at the prediction.

**Table 2:** Overview of the four types of motor sequence quantification for matching. *Abbreviations and variables*: LEMS: lower extremity motor score; MS: motor score; *m*_*i*_ motor score of myotome *i*; NLI: neurological level of injury; *N*: number of myotomes below the NLI, RMSE: root mean squared error; *dist*_*H*_: Hamming distance; *s*_*i*_ score of dermatome or myotome *i* including LTS, PPS, and MS. LTS: light touch score, PPS: pinprick score.

| Match | Description | Formula | Variables used for matching | Pro/Con | Implementation |
| --- | --- | --- | --- | --- | --- |
| Type 1 | The **LEMS** (the sum of the motor scores of the lower extremities) of the patient and any neighbour agrees within $\pm 5$ MS points (increase by 1 point if no match) | $LEMS = \sum_{i=L1}^{S1} m_i$ | LEMS | ⊕ Clinically established score<br>⊕ Simple calculation<br>⊖ Agnostic of the NLI<br>⊖ Agnostic of the MS sequence | *manually in python* |
| Type 2 | The **mean of all MS below the NLI** of patient and any neighbour agrees within $\pm 0.5$ points (increase by 0.1 points if no match) | $meanMS = \frac{1}{N} \sum_{i=NLI}^{S1} m_i$ | MS sequence | ⊕ Accounts for the NLI<br>⊕ Simple calculation<br>⊖ Agnostic of the MS sequence | *manually in python* |
| Type 3 | Patient and any neighbour **MS sequences** below the NLI agree within $\pm 0.5$ points (increase by 0.1 points if no match) quantified by RMSE | $RMSE = \sqrt{\frac{1}{N} \sum_{i=NLI}^{S1} (m_{pat,i} - m_{ref,i})^2}$ | MS sequence | ⊕ Accounts for the NLI<br>⊕ Accounts for the MS sequence<br>⊖ Agnostic of sensory scores | *sklearn.metrics*<br>*mean_squared_error* |
| Type 4A | neighbours are the **k nearest neighbours** by Hamming distance in high dimensional space spanned by all 20 **MSs**. All scores are normalised to the interval [0,1]. k in [1,3,10,20] | $dist_H = \sum_{i=C1}^{S1} d_i$<br>$d_i = \begin{cases} 1 & \text{if } m_{ref,i} = m_{pat,i} \\ 0 & \text{else} \end{cases}$ | MS sequence | ⊕ Accounts for the NLI<br>⊕ Accounts for the MS sequence<br>⊖ Agnostic of sensory score<br>⊖ Fixed number of neighbours | *sklearn.metrics*<br>*pairwise_distance* |
| Type 4B | neighbours are the **k nearest neighbours** by Hamming distance in high dimensional space spanned by all 128 **SSs** and 20 **MSs**. All scores are normalised to the interval [0,1]. k in [1,3,10,20] | $dist_H = \sum_{i=C1}^{S1} d_i$<br>$d_i = \begin{cases} 1 & \text{if } s_{ref,i} = s_{pat,i} \\ 0 & \text{else} \end{cases}$ | MS, LTS, PPS sequences | ⊕ Accounts for the NLI<br>⊕ Accounts for the MS sequence<br>⊕ Accounts for sensory score<br>⊖ Fixed number of neighbours | *sklearn.metrics*<br>*pairwise_distance* |

### 2.3. Statistical analysis and visualisation

#### Motor score assessment uncertainty and bootstrapping

Despite best efforts (32,33) the ISNCSCI scores are subject to uncertainty stemming from the examiner’s level of experience (systematic) and personal fluctuation (random), as well as the patients’ compliance with the examination and natural variation of physical fitness (random variation). We base our estimation of MS uncertainty distributions on a study by Bye *et al*.(14) to calculate the probability density function for each MS level (**Figure S2**). We calculate n=100 bootstraps for each of the target patient’s MS sequences and repeat the matching. In all visualisations we show median values with 95% confidence intervals.

#### Prediction performance reporting

We represent the segmental MS distribution as a graph, whose y-axis shows the MS, and the x-axis the segments of the key muscle from C5 to T1 and L2 to S1 in the order from rostral to caudal. For each patient, we quantify graph agreement based on the root mean squared error below the NLI (RMSE_blNLI_). Models are ranked based on the normalised (to the minimum over all models) sum of the median and 97.5th RMSE_blNLI_ percentiles taken over all patients. The obtained rank is used to identify the best-performing model for neurological recovery prediction.

Functional score prediction performance is quantified as area under the receiver operator characteristic (ROC-AUC) based on the probability of scores of all matched neighbours for a patient.

#### Subgroup assignment and dimensionality reduction

Patient subgroups were assigned using k-means clustering (*sklearn*.*cluster*.*KMeans*) over all acute-phase ISNCSCI scores scaled to [0,1]. We chose a number of eight clusters as a compromise between the minimum number of patients per cluster (99) and subgroup detail covered. We visualise the identified patient subgroups in a 2D-dimensional embedding of the high-dimensional feature space using Uniform Manifold Approximation and Projection (u-map)(34) implemented in the *umap* package.

#### Alternative prediction models

We compare our prediction of walking ability to an unregularized logistic regression (*sklearn*.*metrics*.*LogisticRegression*) based on five input variables: age dichrometised at 65 years, and the larger of the left/right side motor and light touch scores of the gastrocsoleus (S1) and quadriceps femoris (L3) muscles. We performed leave-one-out cross-validation on the EMSCI data, whereas a model trained on all EMSCI data was used for predictions in the Sygen subcohort. Similarly, we provide predictions of MSs at 6 months from a Ridge regression model (*sklearn*.*metrics*.*Ridge*) using all acute-phase MS and SS as inputs. Here hyperparameters were tuned via grid search on the validation subset of a 5-fold nested cross-validation, and results are reported for the pooled test sets over all folds.

## 3. Results

**Figure S1** shows the CONSORT-style flowchart illustrating the selection of 1,267 EMSCI (primary cohort; reference pool) and 411 Sygen (validation cohort) patients. **Table 1** shows relevant summary statistics.

### 3.1. Contribution of reference pool stratification and neighbour agglomeration

**Figure 2A** shows boxplots for the number of identified neighbours given a fixed distance for the similarity metrics Types 1-3 (Type 4 matching yields constant numbers of neighbours) with different options of reference pool stratification. Multiple neighbours, rather than a single match, are frequently identified. Particularly for LEMS and MeanMS similarity (Types 1, 2), a large number of neighbours is identified in the absence of stratification for AIS grade, NLI, and/or sex and age.

**Figure 2.**
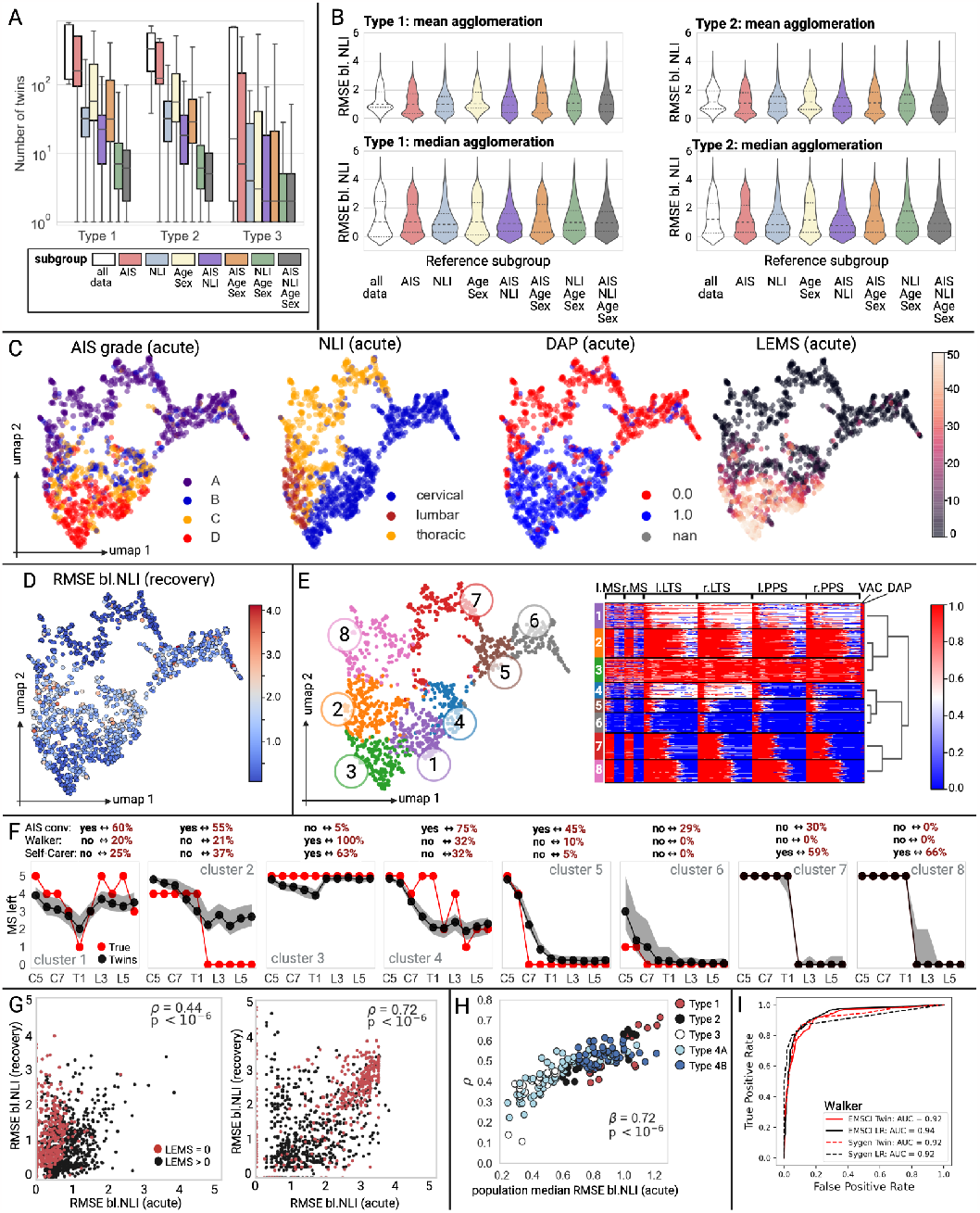
Prediction performance results. A: Boxplots showing the number of neighbours identified for each of the combinations of matching Types 1-3 for the different patient stratifications - i.e. several neighbours are identified for each patient and these are pooled for the final prediction. B: Violin plots of the recovery phase (6 months after SCI) agreement between actual and neighbour-predicted MSs quantified by RMSE_blNLI_. Dotted lines show interquartile ranges, and dashed median values. Results are shown for Type 1 and 2 matching with optional reference patient stratification (hue, x-axis) for mean (top row) and median (bottom row) agglomeration. C: u-map embedding of the included EMSCI cohort with various clinical attributes highlighted. D: The same u-map embedding coloured by RMSE_blNlI_ achieved by the best model. E: u-map embedding coloured by patient cluster (left, numbered 1 to 8) with the corresponding heatmap visualising the contribution of each MS LTS, and PPS to clustering (right). Heatmap colours indicate the level of normalised MS, LTS, or PPS. F: For each cluster, representative examples (achieving the median RMSE_blNLI_ in the relevant cluster) are shown with their true (red) and predicted (black) left-side recovery MSs and functional scores. G: Correlation analysis between the acute and recovery phase RMSE_blNLI_ for the model yielding the best prediction performance for the EMSCI cohort as a whole (left) or the highest Spearman correlation coefficient (LEMS matching with median agglomeration) (right). Each dot indicates one patient. Spearman correlation coefficient (ρ) with p-values (p) are given. H: Summary of all correlation analysis results showing a significant correlation (Spearman correlation coefficient β) between a model’s correlation coefficient ρ and acute phase population median RMSE_blNLI_. Each dot indicates one matching approach. I: ROC with relevant AUCs for the prediction of walking ability for the EMSCI and Sygen patient populations by the historic neighbour or logistic regression. *Abbreviations:* AIS: American Spinal Injury Association impairment scale, NLI: neurological level of injury, LEMS: lower extremity motor score, RMSE_blNLI_: root-mean-squared error below the NLI, u-map: uniform manifold approximation and projection, MS: motor score, LTS: light touch score, PPS: Pinprick score, DAP: deep anal pressure, ROC: receiver operator characteristic, nan: Not a number - missing data for DAP, LR: Logistic regression.

**Figure 2B** shows violin plots of the achieved RMSE_blNLI_ at 6 months between the true segmental MS sequences and the Type 1 and 2 NN predictions (results for other matching approaches and evaluation by ΔLEMS are given in **Figure S3** and yielded minimal variation). Controlling for selected biases prior to NN matching, e.g. by AIS and/or NLI, improved performance (i.e.lower RMSE_blNLI_), whereas the inclusion of age and sex information did not (i.e.comparable shape of the violins to no stratification). Overall, mean agglomeration resulted in narrower distributions, but higher median RMSE_blNLI_ values compared to median agglomeration.

### 3.2. Comparison of similarity metrics

#### Population-level performance

The best combination of reference pool stratification in addition to each of the MS similarity metrics are summarised in **Table 3** (top). For the patient population as a whole, matching Type 4A (20-NN) was best. This model also performed well for AIS grade conversion and functional score predictions (ROC-AUC best for walking ability prediction, 8th for AIS grade conversion, best model without age/sex information for self-care ability prediction). Generally, AIS grade conversion was less accurately predicted and self-care ability prediction benefitted from the inclusion of age and sex information (**Table S1**). Walking ability prediction within the EMSCI cohort (ROC-AUC_kNN_ = 0.92) was comparable to that achieved by a logistic regression model (ROC-AUC_LR(13,35)_= 0.94, **Figure 2I**). Ridge regression achieved an RMSE_blNL_=0.98 (0.22, 2.57) in the same cohort ranking 17th amongst the assessed 176 approaches.

**Table 3:**
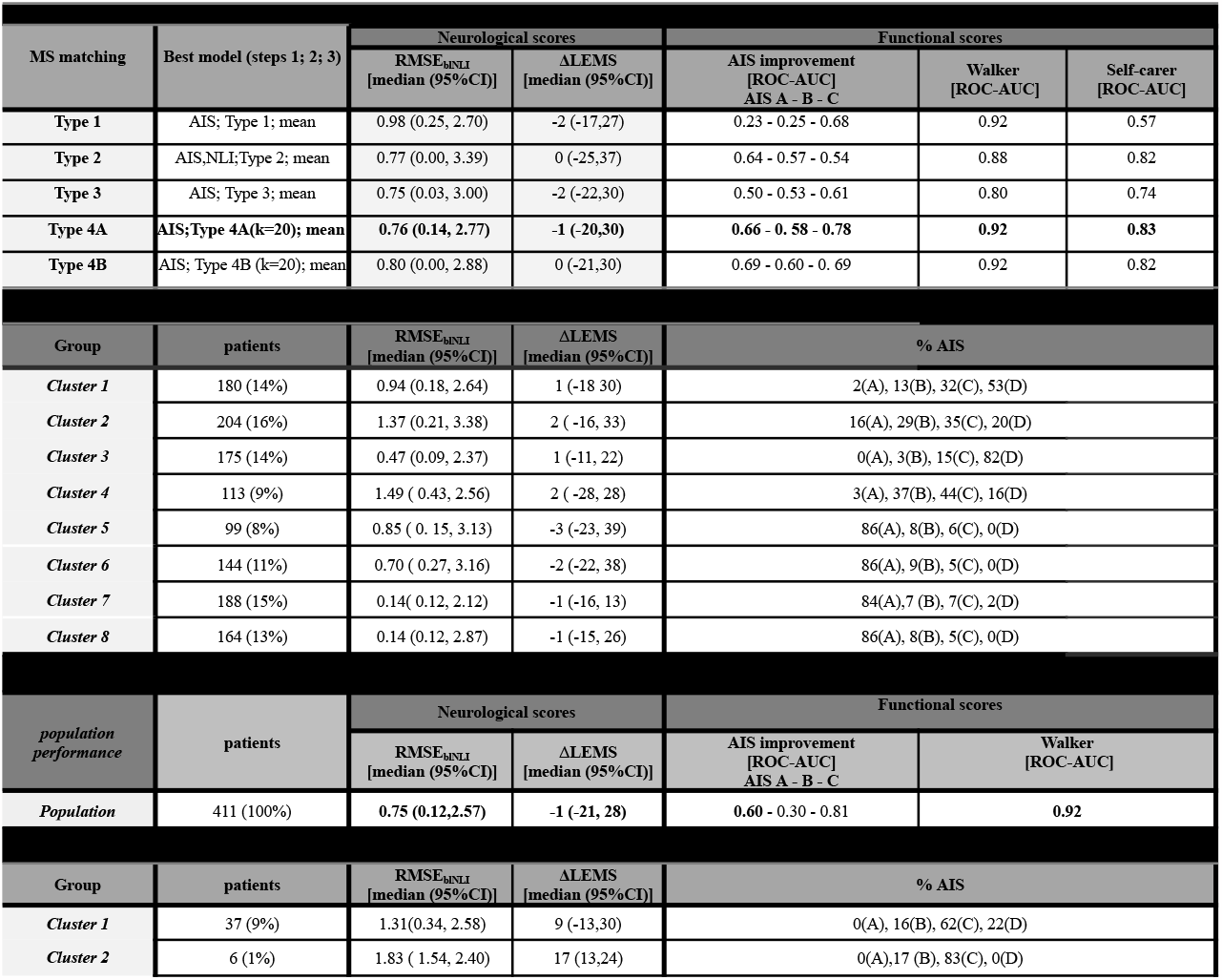
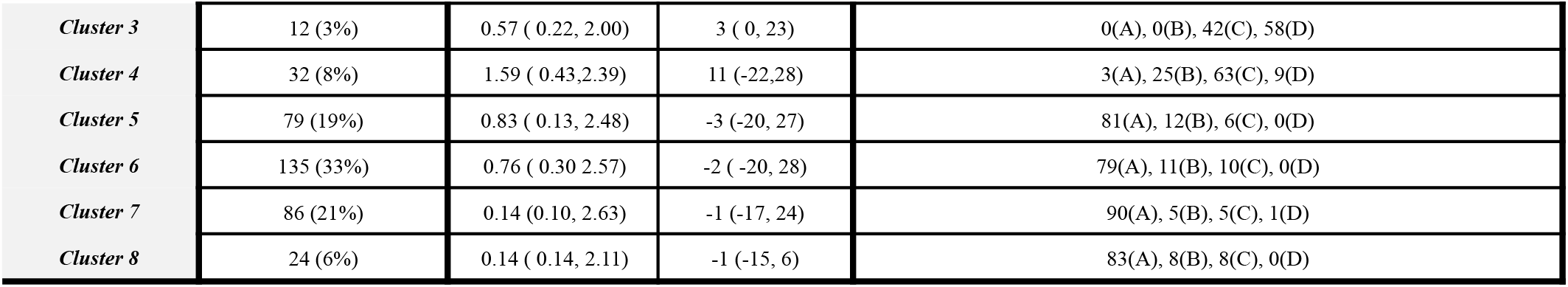
Performance comparison of different neighbour matching approaches evaluated on either the primary or validation cohort. Neurological recovery prediction performance is scored in terms of the RMSE_blNLI_, and ΔLEMS. Functional recovery prediction performance is assessed by AIS grade conversion (by at least one point), walking (SCIM 12 > 4 points), and self-care (SCIM self-care subscore greater than 15 points) ability. Positive class prevalences: EMSCI: AIS A conversion: 0.27, AIS B conversion: 0.66, AIS C conversion: 0.73, walker: 0.35, self-care: 0.57; Validation cohort (Sygen): AIS A conversion: 0.24, AIS B conversion: 0.63, AIS C conversion: 0.87, walker: 0.24. Note that results in clusters comprising very few patients may be subject to large variation (greyed out). *Abbreviations:* AIS: American Spinal Injury Association impairment scale, NLI: neurological level of injury, LEMS: lower extremity motor score, RMSE_blNLI_: root-mean-squared error below the NLI, MS: motor score, ROC-AUC: area under the receiver operator characteristic.

| MS matching | Best model (steps 1; 2; 3) | Neurological scores |  | Functional scores |  |  |
| --- | --- | --- | --- | --- | --- | --- |
| | | $RMSE_{binLI}$<br>[median (95%CI)] | $\Delta LEMS$<br>[median (95%CI)] | AIS improvement<br>[ROC-AUC]<br>AIS A - B - C | Walker<br>[ROC-AUC] | Self-carer<br>[ROC-AUC] |
| Type 1 | AIS; Type 1; mean | 0.98 (0.25, 2.70) | -2 (-17,27) | 0.23 - 0.25 - 0.68 | 0.92 | 0.57 |
| Type 2 | AIS,NLI;Type 2; mean | 0.77 (0.00, 3.39) | 0 (-25,37) | 0.64 - 0.57 - 0.54 | 0.88 | 0.82 |
| Type 3 | AIS; Type 3; mean | 0.75 (0.03, 3.00) | -2 (-22,30) | 0.50 - 0.53 - 0.61 | 0.80 | 0.74 |
| Type 4A | AIS;Type 4A(k=20); mean | 0.76 (0.14, 2.77) | -1 (-20,30) | 0.66 - 0.58 - 0.78 | 0.92 | 0.83 |
| Type 4B | AIS; Type 4B (k=20); mean | 0.80 (0.00, 2.88) | 0 (-21,30) | 0.69 - 0.60 - 0.69 | 0.92 | 0.82 |
| Group | patients | $RMSE_{binLI}$<br>[median (95%CI)] | $\Delta LEMS$<br>[median (95%CI)] | % AIS | | |
| Cluster 1 | 180 (14%) | 0.94 (0.18, 2.64) | 1 (-18 30) | 2(A), 13(B), 32(C), 53(D) |  |  |
| Cluster 2 | 204 (16%) | 1.37 (0.21, 3.38) | 2 (-16, 33) | 16(A), 29(B), 35(C), 20(D) |  |  |
| Cluster 3 | 175 (14%) | 0.47 (0.09, 2.37) | 1 (-11, 22) | 0(A), 3(B), 15(C), 82(D) |  |  |
| Cluster 4 | 113 (9%) | 1.49 ( 0.43, 2.56) | 2 (-28, 28) | 3(A), 37(B), 44(C), 16(D) |  |  |
| Cluster 5 | 99 (8%) | 0.85 ( 0.15, 3.13) | -3 (-23, 39) | 86(A), 8(B), 6(C), 0(D) |  |  |
| Cluster 6 | 144 (11%) | 0.70 ( 0.27, 3.16) | -2 (-22, 38) | 86(A), 9(B), 5(C), 0(D) |  |  |
| Cluster 7 | 188 (15%) | 0.14( 0.12, 2.12) | -1 (-16, 13) | 84(A),7 (B), 7(C), 2(D) |  |  |
| Cluster 8 | 164 (13%) | 0.14 (0.12, 2.87) | -1 (-15, 26) | 86(A), 8(B), 5(C), 0(D) |  |  |
| population performance | patients | Neurological scores |  | Functional scores |  |  |
| | | $RMSE_{binLI}$<br>[median (95%CI)] | $\Delta LEMS$<br>[median (95%CI)] | AIS improvement<br>[ROC-AUC]<br>AIS A - B - C | Walker<br>[ROC-AUC] | |
| Population | 411 (100%) | 0.75 (0.12,2.57) | -1 (-21, 28) | 0.60 - 0.30 - 0.81 | 0.92 |  |
| Group | patients | $RMSE_{binLI}$<br>[median (95%CI)] | $\Delta LEMS$<br>[median (95%CI)] | % AIS | | |
| Cluster 1 | 37 (9%) | 1.31(0.34, 2.58) | 9 (-13,30) | 0(A), 16(B), 62(C), 22(D) |  |  |
| Cluster 2 | 6 (1%) | 1.83 ( 1.54, 2.40) | 17 (13,24) | 0(A),17 (B), 83(C), 0(D) |  |  |

|  |  |  |  |  |
| --- | --- | --- | --- | --- |
| Cluster 3 | 12 (3%) | 0.57 ( 0.22, 2.00) | 3 ( 0, 23) | 0(A), 0(B), 42(C), 58(D) |
| Cluster 4 | 32 (8%) | 1.59 ( 0.43,2.39) | 11 (-22,28) | 3(A), 25(B), 63(C), 9(D) |
| Cluster 5 | 79 (19%) | 0.83 ( 0.13, 2.48) | -3 (-20, 27) | 81(A), 12(B), 6(C), 0(D) |
| Cluster 6 | 135 (33%) | 0.76 ( 0.30 2.57) | -2 ( -20, 28) | 79(A), 11(B), 10(C), 0(D) |
| Cluster 7 | 86 (21%) | 0.14 (0.10, 2.63) | -1 (-17, 24) | 90(A), 5(B), 5(C), 1(D) |
| Cluster 8 | 24 (6%) | 0.14 ( 0.14, 2.11) | -1 (-15, 6) | 83(A), 8(B), 8(C), 0(D) |

#### Performance within patient subgroups

For visualisation, all patient data are projected into a two-dimensional embedding of the full variable spaces using u-map (34) (**Figure 2C)**. By colouring patients in this embedding by selected clinical scores (AIS grade, NLI, LEMS, DAP) subgroups become apparent. **Figure 2D** shows the resulting recovery-phase RMSE_blNLI_ between the true and predicted MSs indicating performance variation as a function of AIS grade and NLI.

We quantify patient subgroup performance in **Table 3** (middle). Subgroups are visualised in **Figure 2E** and separate the cohort by the NLI, AIS grade, and retained motor and sensory function as indicated in the accompanying heatmap. Whereas clusters 1 to 4 comprised more heterogeneous patients, clusters 5-8 included similar AIS A patients as indicated by the degree of noise in the relevant heatmaps and AIS grade composition in **Table 3**. The heatmaps also give an indication of the approximate NLI within the patient subgroups. Within each cluster, some patients did not achieve an acceptable (i.e. RMSE_blNLI_ < 1.0) prediction as indicated by wide 95% confidence intervals of the reported metrics. We observe that clusters comprising a larger fraction of patients with complete (AIS A) paraplegia (clusters 7, 8) or a large proportion of AIS D patients (cluster 3) performed best. Predictions were less accurate for patients with a lumbar injury (cluster 2) or tetraplegic AIS B, and C patients (cluster 4). Representative examples of patients and the 20-NN predictions achieving subgroup median performance scores from each cluster are shown in **Figure 2F**. All predictions are provided with 95% confidence bands for MSs, and a probability in addition to a binary label for functional endpoints. This illustration gives a notion of the clinical relevance of the achieved median RMSE_blNLI_ given uncertainty in the ISNCSCI score assessment. Independent of the chosen similarity metric, and patient subpopulation, we observe substantial variation in prediction performance by patients, reflected by large ranges of the evaluation metrics.

### 3.3. Correlation analysis between acute and recovery phase neighbour agreement

One possible explanation for poor prediction is suboptimal NN matching in the acute phase motivating a correlation analysis of the acute and recovery phase RMSE_blNL_ between the agglomerated neighbour cohorts and patient of interest. **Figure 2G** shows two representative examples of this analysis, the matching performing best for the EMSCI cohort (**Figure 2G**, left), and the model achieving the highest correlation coefficient (no added stratification; Type 1; median) in **Figure 2G**, right. We observe that Type 1 matching may not always achieve good agreement between the neighbours’ and the acute phase segmental MSs (RMSE_blNLI_ > 2), particularly for patients with an acute phase LEMS of 0 likely due to not accounting for impairment of the upper extremities. These patients could also not be predicted well, even if matched perfectly in the acute phase (see **Figure 2G**, right). In contrast, good matches were identified for most patients in the acute phase for Type 4A kNN but this did not always translate to an accurate prediction, i.e. low RMSE_blNLI_ at 6 months (**Figure 2G**, left). In general, independent of the similarity and evaluation metric, all correlations were highly significant (all p-values < 0.0001), while correlation coefficients varied markedly as summarised in **Figure 2H**. The highest correlation coefficients were observed for similarity assessed by LEMS and MeanMS (Type 1 and 2). A stronger correlation implies that if a good/poor match is identified, a reasonable/poor recovery prediction is more likely. We observe that stronger correlation coefficients are driven by poorer matching in the acute phase, leading also to poorer prediction. Only 62% of patients displayed a very good match (RMSE_blNLI_ < 0.3) to their identified nearest neighbour cohort in the acute phase for any tested model. More severely injured patients were generally matched better: AIS A: 490/546 (90%), B: 154/178 (87%), C: 80/239 (33%), D: 61/304 (20%). The fraction of patients achieving an acceptable prediction (RMSE_blNLI_ ≦ 0.5) also varied greatly depending on the used matching method. They ranged between 99 (subset by age, sex, and NLI with 20-NN(MS, SS), mean agglomeration) to 512 (no subset, 20-NN(MS), median aggregation) of the 1,267 EMSCI patients. Specifically, matching approaches relying on stratification by age and sex resulted in fewer acceptable matches. For models based on MS (i.e., not accounting for sensory scores), neighbour agglomeration by median led to stronger correlations than mean averaging. Here, we also observed that correlation coefficients increased (associated p-values decreased) with an increasing number of neighbours considered for Type 4 (kNN) matching.

### 3.4. Validation cohort

We tested the proposed pipeline on an independent cohort derived from the Sygen trial (**Table 3**, bottom). Here, the same model identified for the EMSCI cohort (AIS; Type 4 (k=20); mean) performed best in terms of neurological recovery and achieved a comparable ROC-AUC for walking ability prediction as in the primary cohort and a logistic regression model (**Figure 2I**, ROC-AUC_LR_ = ROC-AUC_kNN Sygen_ = 0.92). Results in terms of RMSE_blNLI_ and ΔLEMS were also comparable for patient subgroups and the population as a whole (**Table 3**, middle and bottom) with the worst performance observed in clusters 2 and 4, whereas patients in clusters 7 or 8 had the best prediction performance.

## 4. Discussion

kNN has previously been applied in the context of SCI predictions but outside the realm of recovery prediction from ISNCSCI scores (36–38). Given its ease of calculation and direct interpretability, kNN is particularly promising in the medical sector for highly heterogeneous, yet small patient cohorts characterised by few clinical variables, such as SCI. The key findings of this study include i) the identification of an optimal historic cohort matching strategy, ii) the observation that multiple rather than single reference patient provide superior predictions, and iii) competitive prediction performance on two large cohorts for neurological and functional endpoints with prediction confidence.

i. We investigated several similarity metrics aiming for robust (independent validation), highly-detailed (prediction of full motor function pattern), and diverse (functional and neurological) recovery predictions. This goes beyond previous work (39,40) in all of these aspects. Historic cohorts identified by kNN are versatile and address several aspects of the recovery process. Based on ISNCSCI scores kNN outperformed linear regression for the prediction of motor function patterns and achieved comparable performance for functional endpoints as supervised logistic regression. Expectedly, subgroup analysis showed that patients presenting with either severe (AIS A) or mild injuries (AIS D) achieved better predictive performance. For AIS A patients, this may be due to a larger number of patients (EMSCI: 43%, Sygen: 65%), better matching performance in the acute injury phase (i.e. significant correlation of early and recovery-phase RMSE_blNLI_), and an overall smaller recovery achieved (flooring effect). In contrast, ceiling effects may motivate AIS D performance. Similar to previous studies, we recorded the strongest variation for AIS B and C patients, which represent a small fraction of the included patients (∼30%) but display large variations in motor and sensory function.
ii. 20-NN assigns *a historic cohort* rather than a single, closest neighbour yielding more generalizable predictions than 1NN. This aligns with the notion that no two patients recover the same but rather that similar patterns of recovery exist. For neurological recovery prediction it helped to stratify the reference pool by AIS grade but no other demographic or injury characteristic in addition to the MS quantification. In agreement with previous work (28), age or sex infomration did not improve neurological outcome predictions. MSs are intrinsically reported relative to the expected maximum possible implying a potential assessment bias with respect to age and sex. However, we observed that age and sex information benefitted the prediction of self-care ability.
iii. Several studies have previously addressed the challenge of SCI recovery prediction of various endpoints such as walking scores, AIS grade conversion, or total motor score recovery.(17,35,41,42) Despite promising results within small cohorts, including detailed additional data such as cerebrospinal fluid biomarkers or imaging data, (11,43) a more generalizable approach based only on clinically well-established scores allowing for large cohort evaluations was missing. Our analysis fills this gap with a data-driven, personalised, and multi-faceted recovery prediction for the individual. For walking ability, we achieve comparable performance as previously reported based on a logistic regression model (13,35), however, our approach provides a variety of additional endpoint predictions and each of these with confidence estimates. As such, we improved beyond the state-of-the-art in terms of prediction detail and interpretability of the underlying model. The evaluation of functional endpoints was key in addition to quantifying neurological recovery here. ΔLEMS or RMSE_blNLI_ are not directly related to clinical outcomes making it difficult to interpret the clinical relevance of the prediction performance in terms of these scores. Still, our study presents quantitative results for these metrics that serve as a baseline for future prediction models. The clinical impact of this study is twofold. Firstly, the presented concept provides patients and physicians with a personalised perspective regarding neurological and functional recovery, including quantification of prediction confidence stemming from the distribution in the reference cohort. The prediction process is inherently interpretable by reviewing the selected neighbours. Moreover, we analysed patient subgrouping through clustering beyond the current gold standard AIS grades alone which allows for more fine-grained and noise-robust group assignments also for other clinical evaluations. Secondly, we provide a means to identify historic controls for individual trial patients to increase and improve the pool of controls in clinical trials. This is of particular importance given the large heterogeneity and low incidence of SCI. Our publicly available code enables direct application of the concept. An *in silico* evaluation of the statistical power of this way to identify historic control cohorts will be addressed in our follow-up study.

Some limitations remain. kNN performance varied among patient subgroups, particularly those reflecting the highest degree of heterogeneity in terms of injury and recovery potential. Also, despite very good performance for walking ability and self-caring, AIS grade conversion was poorly predicted for AIS A patients. This is likely due to low incidence of improving AIS A patients and implies that the underlying mechanisms are not reflected in the ISNCSCI exam. Instead, factors not included, e.g. CSF, imaging, or other modifying events may drive this outcome. The data included lacked potentially important clinical information, e.g. the timing and success of decompression surgery, patient comorbidities, or other treatment and response effects (e.g. blood markers, medication). Such information was not available for a sufficiently large patient population in a database. Our approach, based on the ISNCSCI exam, was a compromise between the number of patients and the assessment detail covered. We suggest repeating the presented evaluation once more detailed data become available.

We further did not account for longitudinal variation during matching. Adverse, disease-modifying events, e.g. severe pneumonia, may greatly influence recovery (44). Since we restricted this analysis to a single time point for neighbour matching, we ignored longitudinal changes in recovery trajectory due to modifying events and excluded patients that greatly deteriorated. However, a lot of the observed variation in the AIS B and C categories may be driven by modifying events. The proposed analysis hence represents predictions based on an expected normal/optimal recovery trajectory. Given a more detailed data pool, it would be possible to consider matching at multiple longitudinal time points. We purposefully did not include longitudinal information of later time points as the aim of a historic cohort by our interpretation should be i) a perspective of recovery in the very acute injury phase, and ii) historic control cohort building for trials in which patients are randomised in the acute injury phase.

Patient numbers and the comparably early time of evaluation (6 months) may be a further limitation. The EMSCI database is one of the largest, longitudinal SCI databases to date assessing patients at 26 and 52 weeks at the latest. Owing to large proportions of missing data, it was not possible to include all enrolled patients and motivated the use of the 6-month time point given larger data availability. Our kNN is strongly based on motor and sensory scores which are difficult to impute leading to complete-case analysis, i.e. discarding any patients with missing data (EMSCI 75 %, Sygen 46%, see **Figure S1**). We would like to stress that SCI recovery at 6 months is likely not final and further improvements are possible. However, we presented a proof-of-principle evaluation for a single time point (six months) that is translatable to different evaluation endpoints.

Another limitation of the concept in itself is the lack of any underlying mechanistic modeling or description of potential relations among variables and predictions as is the case for other machine learning and mathematical models. Thus, this approach cannot infer beyond the provided pool of reference patients whereas other algorithms may increase inference strength given a sufficiently large number of training examples. This especially limits performance for patient subgroups where variation in motor and sensory scores is large, but sample numbers are limited (here AIS B and C). It would be essential to increase the pool of historic reference patients to boost the performance of kNN for the purpose of clinical application but also to allow for data-driven extrapolation and pattern mining to improve predictions. Despite investigating several similarity metrics, it is clear that this is not a fully exhaustive evaluation, and further optimization of the matching criteria, e.g. of the LEMS or meanMS matching thresholds may lead to different results for these matching types.

## 5. Conclusions

We presented a systematic analysis of the concept of nearest neighbour matching for recovery prediction of traumatic SCI based on acute injury phase ISNCSCI assessment scores. We further propose quantitative metrics to compare prediction performance and apply these to identify the optimal matching criteria for specific patient subgroups. By making our software pipeline publicly available and validating our findings in an independent test cohort, we provide a path of clinical translation of the proposed concept in terms of cohort design and evaluation of clinical trials in the realm of SCI research in addition to providing personalised recovery predictions.

## Data Availability

All data produced in the present study are available upon reasonable request to the authors.

## Declarations

### Ethics approval and consent to participate

The EMSCI study is performed in accordance with the Declaration of Helsinki and approved by all contributing institutional review boards. EMSCI follows the ethical guidelines of the participating countries and patients gave their written informed consent before being included in the database. The Sygen clinical trial also received ethical approval but was conducted before clinical trials were required to be registered (i.e., no clinicaltrial.gov identifier available).

### Consent for publication

Not applicable.

### Availability of data and materials

Anonymized data used in this study will be made available upon request to the corresponding author and in compliance with the General Data Protection Regulation (EU GDPR). We publish all code required to reproduce the presented results in our GitHub repository: https://github.com/BorgwardtLab/SCIRecoveryPredictionPublic.git

### Authors’ contribution

SB implemented the presented models and performed the evaluation, visualisation, and interpretation of the results. LB and LL supported the design of the matching algorithms and patient cohorts. The EMSCI Study group collected data used in this analysis. AC led data collection and provided clinical insight to the prediction task. DM, RA, NW, RR, and JG gave critical feedback to improve the conceptual design and supported the clinical interpretation of the proposed findings. CJ designed the study and provided data access and continuous feedback. All authors contributed to the writing of the manuscript and reviewed the written document.

## List of abbreviations

AIS: American Spinal Injury Association impairment scale
ASIA: The American Spinal Injury Association
DAP: Deep anal pressure
EMSCI: European Multicenter Study about Spinal Cord Injury
ISNCSCI: International Standards for Neurological Classification of SCI
kNN: k-nearest neighbour
LEMS: Lower extremity motor score
LR: Logistic regression
LTS: Light touch score
meanMS: Mean motor score
MS: Motor score
nan: Not a number - missing data for DAP
NLI: Neurological level of injury
PPS: Pinprick score
RMSE_blNLI_: Root-mean-squared error below the NLI
ROC: Receiver operator characteristic
SCI: Spinal cord injury
SCIM: Spinal cord independence measure.

## References

1. Badhiwala JH, Wilson JR, Fehlings MG. Global burden of traumatic brain and spinal cord injury. Lancet Neurol. 2019 Jan;18(1):24–5.

2. Post M, Noreau L. Quality of life after spinal cord injury. J Neurol Phys Ther. 2005 Sep;29(3):139–46.

3. Fawcett JW, Curt A, Steeves JD, Coleman WP, Tuszynski MH, Lammertse D, et al. Guidelines for the conduct of clinical trials for spinal cord injury as developed by the ICCP panel: spontaneous recovery after spinal cord injury and statistical power needed for therapeutic clinical trials. Spinal Cord. 2007 Mar;45(3):190–205.

4. Kramer JLK, Lammertse DP, Schubert M, Curt A, Steeves JD. Relationship between motor recovery and independence after sensorimotor-complete cervical spinal cord injury. Neurorehabil Neural Repair. 2012 Nov;26(9):1064–71.

5. DeVries Z, Hoda M, Rivers CS, Maher A, Wai E, Moravek D, et al. Development of an unsupervised machine learning algorithm for the prognostication of walking ability in spinal cord injury patients. Spine J. 2020 Feb;20(2):213–24.

6. Wilson JR, Grossman RG, Frankowski RF, Kiss A, Davis AM, Kulkarni AV, et al. A clinical prediction model for long-term functional outcome after traumatic spinal cord injury based on acute clinical and imaging factors. J Neurotrauma. 2012 Sep;29(13):2263–71.

7. Kaminski L, Cordemans V, Cernat E, M’Bra KI, Mac-Thiong JM. Functional Outcome Prediction after Traumatic Spinal Cord Injury Based on Acute Clinical Factors. J Neurotrauma. 2017 Jun 15;34(12):2027–33.

8. Kirshblum S, Snider B, Eren F, Guest J. Characterizing Natural Recovery after Traumatic Spinal Cord Injury. J Neurotrauma. 2021 May 1;38(9):1267–84.

9. Kirshblum SC, Botticello AL, Dyson-Hudson TA, Byrne R, Marino RJ, Lammertse DP. Patterns of Sacral Sparing Components on Neurologic Recovery in Newly Injured Persons With Traumatic Spinal Cord Injury. Arch Phys Med Rehabil. 2016 Oct;97(10):1647–55.

10. Wilson JR, Jaja BNR, Kwon BK, Guest JD, Harrop JS, Aarabi B, et al. Natural History, Predictors of Outcome, and Effects of Treatment in Thoracic Spinal Cord Injury: A Multi-Center Cohort Study from the North American Clinical Trials Network [Internet]. Vol. 35, Journal of Neurotrauma. 2018. p. 2554–60. Available from: 10.1089/neu.2017.5535

11. Freund P, Seif M, Weiskopf N, Friston K, Fehlings MG, Thompson AJ, et al. MRI in traumatic spinal cord injury: from clinical assessment to neuroimaging biomarkers. Lancet Neurol. 2019 Dec;18(12):1123–35.

12. Dalkilic T, Fallah N, Noonan VK, Salimi Elizei S, Dong K, Belanger L, et al. Predicting Injury Severity and Neurological Recovery after Acute Cervical Spinal Cord Injury: A Comparison of Cerebrospinal Fluid and Magnetic Resonance Imaging Biomarkers. J Neurotrauma. 2018 Feb 1;35(3):435–45.

13. van Middendorp JJ, Hosman AJF, Donders ART, Pouw MH, Ditunno JF Jr, Curt A, et al. A clinical prediction rule for ambulation outcomes after traumatic spinal cord injury: a longitudinal cohort study. Lancet. 2011 Mar 19;377(9770):1004–10.

14. Bye E, Glinsky J, Yeomans J, Hungerford A, Patterson H, Chen L, et al. The inter-rater reliability of the 13-point manual muscle test in people with spinal cord injury. Physiother Theory Pract. 2021 Oct;37(10):1126–31.

15. Feigin VL, Nichols E, Alam T, Bannick MS, Beghi E, Blake N, et al. Global, regional, and national burden of neurological disorders, 1990–2016: a systematic analysis for the Global Burden of Disease Study 2016. Lancet Neurol. 2019 May 1;18(5):459–80.

16. Kucher K, Johns D, Maier D, Abel R, Badke A, Baron H, et al. First-in-Man Intrathecal Application of Neurite Growth-Promoting Anti-Nogo-A Antibodies in Acute Spinal Cord Injury. Neurorehabil Neural Repair. 2018 Jun;32(6-7):578–89.

17. Belliveau T, Jette AM, Seetharama S, Axt J, Rosenblum D, Larose D, et al. Developing Artificial Neural Network Models to Predict Functioning One Year After Traumatic Spinal Cord Injury. Arch Phys Med Rehabil. 2016 Oct;97(10):1663–8.e3.

18. Khan O, Badhiwala JH, Wilson JRF, Jiang F, Martin AR, Fehlings MG. Predictive Modeling of Outcomes After Traumatic and Nontraumatic Spinal Cord Injury Using Machine Learning: Review of Current Progress and Future Directions. Neurospine. 2019 Dec;16(4):678–85.

19. Inoue T, Ichikawa D, Ueno T, Cheong M, Inoue T, Whetstone WD, et al. XGBoost, a Machine Learning Method, Predicts Neurological Recovery in Patients with Cervical Spinal Cord Injury. Neurotrauma Rep. 2020 Jul 23;1(1):8–16.

20. McCoy DB, Dupont SM, Gros C, Cohen-Adad J, Huie RJ, Ferguson A, et al. Convolutional Neural Network-Based Automated Segmentation of the Spinal Cord and Contusion Injury: Deep Learning Biomarker Correlates of Motor Impairment in Acute Spinal Cord Injury. AJNR Am J Neuroradiol. 2019 Apr;40(4):737–44.

21. k-nearest neighbor algorithm. In: Discovering Knowledge in Data. Hoboken, NJ, USA: John Wiley & Sons, Inc.; 2014. p. 149–64.

22. Geisler FH, Coleman WP, Grieco G, Poonian D, Sygen Study Group. The Sygen multicenter acute spinal cord injury study. Spine. 2001 Dec 15;26(24 Suppl):S87–98.

23. Kirshblum SC, Burns SP, Biering-Sorensen F, Donovan W, Graves DE, Jha A, et al. International standards for neurological classification of spinal cord injury (revised 2011). J Spinal Cord Med. 2011 Nov;34(6):535–46.

24. Rupp R, Biering-Sørensen F, Burns SP, Graves DE, Guest J, Jones L, et al. International Standards for Neurological Classification of Spinal Cord Injury: Revised 2019. Top Spinal Cord Inj Rehabil. 2021 Spring;27(2):1–22.

25. Geisler FH, Coleman WP, Grieco G, Poonian D, Sygen Study Group. Recruitment and early treatment in a multicenter study of acute spinal cord injury. Spine. 2001 Dec 15;26(24 Suppl):S58–67.

26. Cornea CM, Silva NA, Marble WS, Hooten K, Sindelar B. Evolution of spinal cord injury treatment in military neurosurgery. Neurosurg Focus. 2022 Sep;53(3):E11.

27. Schiller MD, Mobbs RJ. The historical evolution of the management of spinal cord injury. J Clin Neurosci. 2012 Oct;19(10):1348–53.

28. Bourguignon L, Tong B, Geisler F, Schubert M, Röhrich F, Saur M, et al. International surveillance study in acute spinal cord injury confirms viability of multinational clinical trials. BMC Med. 2022 Jun 14;20(1):225.

29. Catz A, Itzkovich M, Steinberg F, Philo O, Ring H, Ronen J, et al. The Catz-Itzkovich SCIM: a revised version of the Spinal Cord Independence Measure. Disabil Rehabil. 2001 Apr 15;23(6):263–8.

30. Itzkovich M, Shefler H, Front L, Gur-Pollack R, Elkayam K, Bluvshtein V, et al. SCIM III (Spinal Cord Independence Measure version III): reliability of assessment by interview and comparison with assessment by observation. Spinal Cord. 2018 Jan;56(1):46–51.

31. Benzel EC, Lancon J, Kesterson L, Hadden T. Cervical laminectomy and dentate ligament section for cervical spondylotic myelopathy. J Spinal Disord. 1991 Sep;4(3):286–95.

32. Franz S, Heutehaus L, Weinand S, Weidner N, Rupp R, Schuld C. Theoretical and practical training improves knowledge of the examination guidelines of the International Standards for Neurological Classification of Spinal Cord Injury. Spinal Cord. 2022 Jan;60(1):1–10.

33. Schuld C, Wiese J, Franz S, Putz C, Stierle I, Smoor I, et al. Effect of formal training in scaling, scoring and classification of the International Standards for Neurological Classification of Spinal Cord Injury. Spinal Cord. 2013 Apr;51(4):282–8.

34. McInnes L, Healy J, Melville J. UMAP: Uniform Manifold Approximation and Projection for Dimension Reduction [Internet]. arXiv [stat.ML]. 2018. Available from: http://arxiv.org/abs/1802.03426

35. Hicks KE, Zhao Y, Fallah N, Rivers CS, Noonan VK, Plashkes T, et al. A simplified clinical prediction rule for prognosticating independent walking after spinal cord injury: a prospective study from a Canadian multicenter spinal cord injury registry. Spine J. 2017 Oct;17(10):1383–92.

36. Popp WL, Schneider S, Bär J, Bösch P, Spengler CM, Gassert R, et al. Wearable Sensors in Ambulatory Individuals With a Spinal Cord Injury: From Energy Expenditure Estimation to Activity Recommendations. Front Neurol. 2019 Nov 1;10:1092.

37. Masood F, Sharma M, Mand D, Nesathurai S, Simmons HA, Brunner K, et al. A Novel Application of Deep Learning (Convolutional Neural Network) for Traumatic Spinal Cord Injury Classification Using Automatically Learned Features of EMG Signal. Sensors [Internet]. 2022 Nov 3;22(21). Available from: 10.3390/s22218455

38. Kayikcioglu T, Aydemir O. A polynomial fitting and k-NN based approach for improving classification of motor imagery BCI data. Pattern Recognit Lett. 2010 Aug 1;31(11):1207–15.

39. Zörner B, Blanckenhorn WU, Dietz V, EM-SCI Study Group, Curt A. Clinical algorithm for improved prediction of ambulation and patient stratification after incomplete spinal cord injury. J Neurotrauma. 2010 Jan;27(1):241–52.

40. Lee BA, Leiby BE, Marino RJ. Neurological and functional recovery after thoracic spinal cord injury. J Spinal Cord Med. 2016;39(1):67–76.

41. Chay W, Kirshblum S. Predicting Outcomes After Spinal Cord Injury. Phys Med Rehabil Clin N Am. 2020 Aug;31(3):331–43.

42. Denis AR, Feldman D, Thompson C, Mac-Thiong JM. Prediction of functional recovery six months following traumatic spinal cord injury during acute care hospitalization. J Spinal Cord Med. 2018 May;41(3):309–17.

43. Kim JH, Song SK, Burke DA, Magnuson DSK. Comprehensive locomotor outcomes correlate to hyperacute diffusion tensor measures after spinal cord injury in the adult rat. Exp Neurol. 2012 May;235(1):188–96.

44. Kopp MA, Watzlawick R, Martus P, Failli V, Finkenstaedt FW, Chen Y, et al. Long-term functional outcome in patients with acquired infections after acute spinal cord injury. Neurology. 2017 Feb 28;88(9):892–900.

