## Supplementary material for "An unsupervised machine learning approach to predict recovery from traumatic spinal cord injury"

**Figure S1:** CONSORT flow-chart indicating the included and excluded patients from the EMSCI, and the Sygen trial. We limited this evaluation to patients with complete ISNCSCI assessments of motor and sensory functions, including AIS grade at the acute injury phase and 26 weeks after recovery. Moreover, we excluded patients that showed significant deterioration in their motor scores, indicated by a decrease of more than one point between acute and 6 months. As such deviations are highly unlikely to be due to assessment uncertainty, but rather a consequence of an adverse disease-modifying intercurrent event (e.g. severe pneumonia), such behaviour would not be captured by historic twin matching in the absence of detailed information.
*Abbreviations:* AIS: American Spinal Injury Association impairment scale, NLI: neurological level of injury; MS: motor score

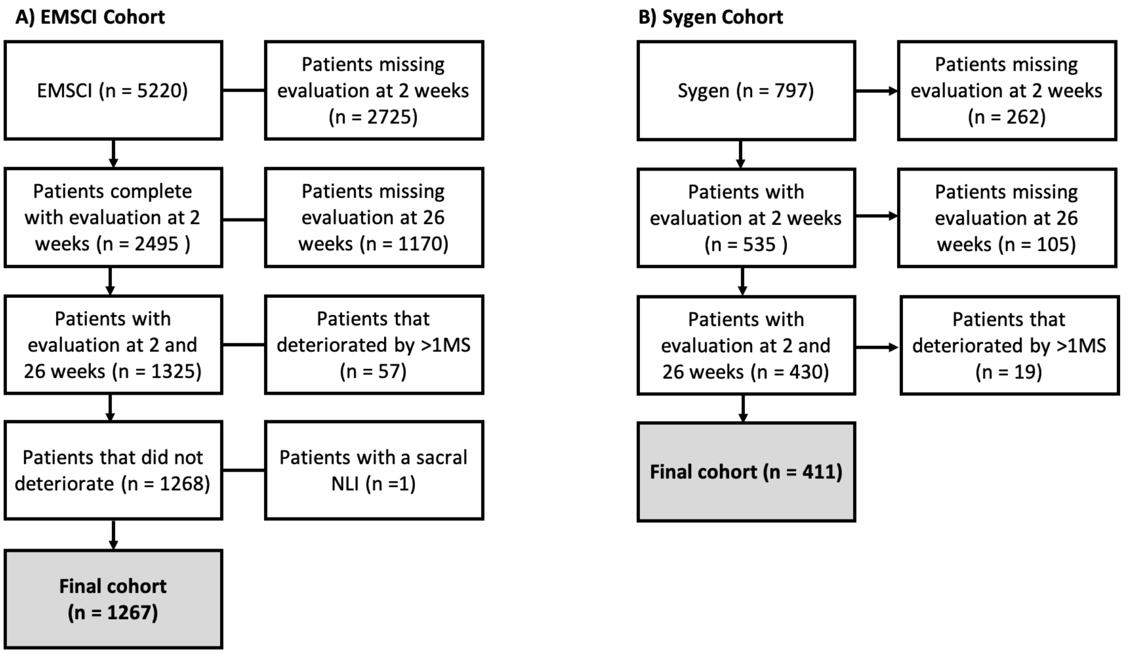


**Figure S2:** Probability density function of the motor assessment uncertainty following an analysis of Bye et al. [38]. Bars are shown for each of the five motor scores (MS) with relevant contributions to assessment levels. The true score of an assessment was assigned by a majority vote.

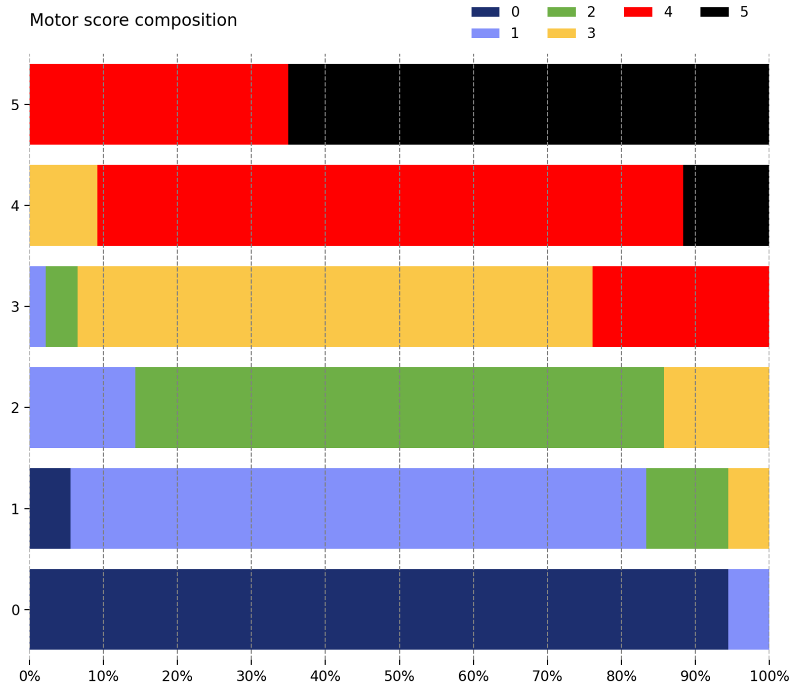

**Figure S3** Violin plots of the recovery phase agreement between true and twin-predicted MS sequences quantified by $RMSE_{blNLI}$ or ΔLEMS for different matching types. Lines show interquartile ranges. Models are grouped by MS matching metric with optional patient subgrouping (hue, x-axis). Results are shown for either mean or median averaging as indicated at the column heads.
*Abbreviations:* AIS: American Spinal Injury Association impairment scale, NLI: neurological level of injury, LEMS: lower extremity motor score, $RMSE_{blNLI}$: root-mean-squared error below the NLI

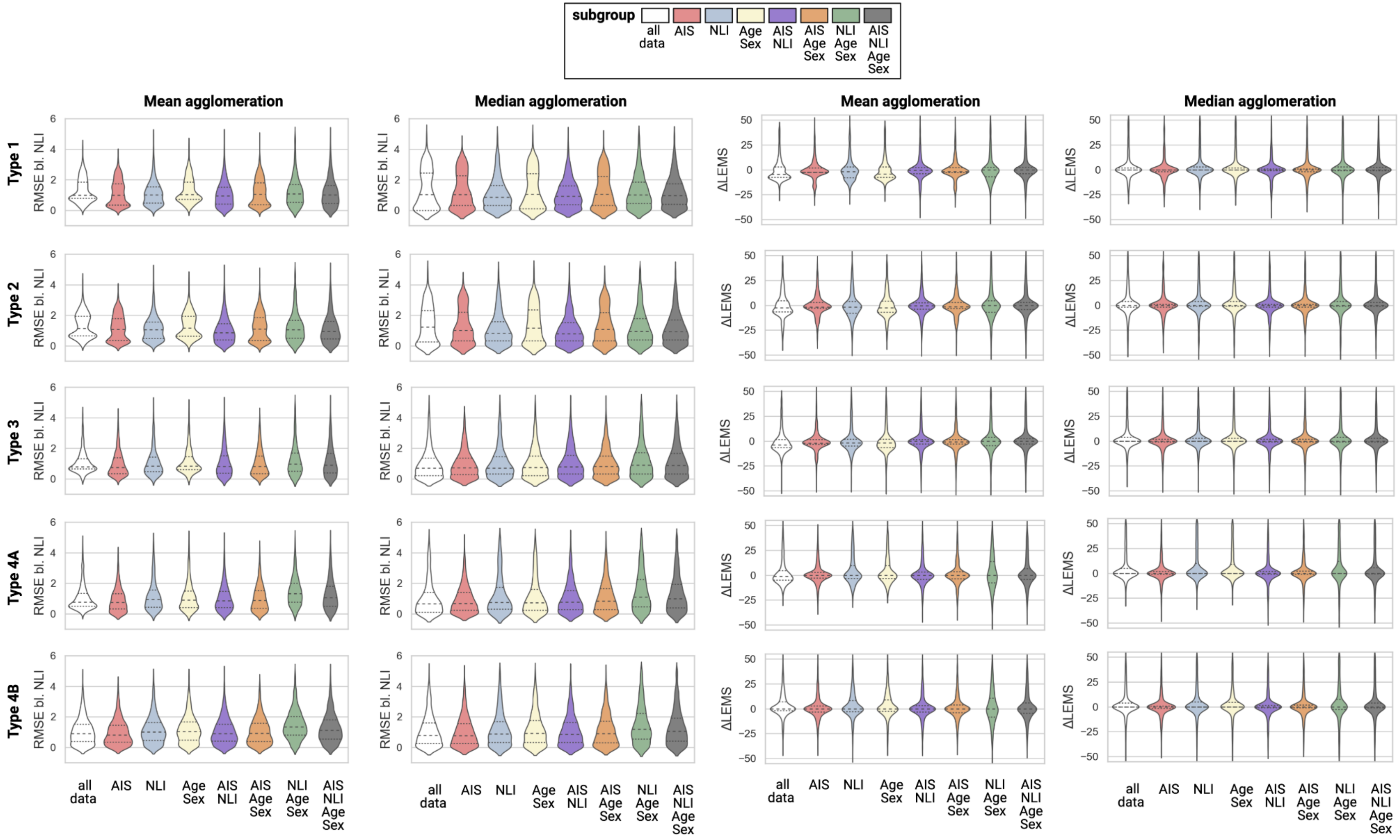

**Table S1:** Functional score prediction results on the EMSCI cohort for each of the 176 models (uniform application), as well as cluster and individual-based matching. Predicted scores include the ROC-AUC for the individual AIS grades (A-E) in a one-vs-rest setting, ROC-AUC values for AIS grade conversion of AIS grade A, B, and C, as well as for independent walking and self-care ability.
*Abbreviations:* AIS: American Spinal Injury Association impairment scale, ROC-AUC: area under the receiver operator characteristic.

**Table S2:** Functional score prediction results on the Sygen cohort for each of the 176 models (uniform application), as well as cluster and individual-based matching. Predicted scores include the ROC-AUC for the individual AIS grades (A-E) in a one-vs-rest setting, ROC-AUC values for AIS grade conversion of AIS grade A, B, and C, as well as for independent walking ability.
*Abbreviations:* AIS: American Spinal Injury Association impairment scale, ROC-AUC: area under the receiver operator characteristic.
